# Epidemic size of novel coronavirus-infected pneumonia in the Epicenter Wuhan: using data of five-countries’ evacuation action

**DOI:** 10.1101/2020.02.12.20022285

**Authors:** Hongxin Zhao, Sailimai Man, Bo Wang, Yi Ning

## Abstract

**Background:** Since late December 2019, novel coronavirus–infected pneumonia (NCP) emerged in Wuhan, Hubei province, China. Meanwhile, NCP rapidly spread from China to other countries, and several countries’ government rush to evacuate their citizens from Wuhan. We analyzed the infection rate of the evacuees and extrapolated the results in Wuhan’s NCP incidence estimation.

**Methods:** We collected the total number and confirmed cases of 2019-nCov infection in the evacuation of Korea, Japan, Germany, Singapore, and France and estimated the infection rate of the 2019 novel coronavirus (2019-nCov) among people who were evacuated from Wuhan with a meta-analysis. NCP incidence of Wuhan was indirectly estimated based on data of evacuation.

**Results:** From Jan 29 to Feb 2, 2020, 1916 people have been evacuated from Wuhan, among them 17 have been confirmed 2019-nCov infected. The infection rate is estimated to be 1.1% (95% CI 0.4%-3.1%) using one group meta-analysis method with random effect model. We then estimated that almost 110,000 (95% CI: 40,000-310,000) people were infected with 2019-nCov in Wuhan around Feb 2, 2020, assuming the infection risk of evacuees is close to Chinese citizens in Wuhan.

**Conclusions:** At the beginning of the outbreak, incidence of NCP may be vastly underestimated. Our result emphasizes that 2019-nCov has proposed a huge public health threats in Wuhan. We need to respond more rapidly, take large-scale public health interventions and draconian measures to limiting population mobility and control the epidemic.

## Introduction

In the late December 2019, a pneumonia, now known as novel coronavirus–infected pneumonia (NCP) caused by the 2019 novel coronavirus (2019-nCoV), emerged in Wuhan, Hubei province, China ^[1]^. The outbreak of NCP quickly showed the characteristic of human-to-human transmission. Chinese health authorities did an immediate investigation to characterize and control the disease, including isolation of suspected cases, monitoring of close contacts, collection of epidemiological and clinical data from confirmed patients, and development of diagnostic and treatment procedures ^[2]^. As of Feb 4, 2020, a total of 24,324 cases with laboratory-confirmed 2019-nCoV infection have been detected in China, of whom 490 have died ^[3]^. There were 8,351 cases confirmed in Wuhan, 362 of the confirmed cases have died, the case fatality was much higher in Wuhan than in the other areas of China ^[4]^. Meanwhile, NCP rapidly spread from China to other countries. After assessing the virus-related information, the WHO decided to declare it as a public health emergency of international concern (PHEIC) on Jan 31, 2020 ^[5]^.

Consequently, to reduce further spread of the 2019-nCoV within China and other countries, remarkable public health measures have been taken and implemented in China. The local government in Wuhan announced on Jan 23, 2020, the suspension of public transportation, with closure of airports, railway stations, and highways in the city, to prevent further disease transmission ^[6]^. Wuhan as the epicenter of this outbreak, the outbreak size is of great significance for understanding the status of the epidemic in Wuhan, predicting the trend of the epidemic, and formulating strategy for prevention and control. Several studies estimated the outbreak size in Wuhan to date. Using a transmission model, British researchers estimated Wuhan case ascertainment of 5.0% (3.6-7.4); 21,022 (11,090-33,490) total infections in Wuhan from 1 to 22 Jan 2020 ^[7]^. In addition, researchers from the University of Hong Kong, China, based on data of confirmed cases published by the Chinese Center for Disease Control and Prevention, and data of domestic and international travel, estimated 75,815 individuals (95% CI 37,304-130,330) have been infected in Wuhan as of Jan 25, 2020. Although the results are significantly higher compared to the figures published officially in Wuhan, the researchers still believed there may be possibilities that they have underestimated the outbreak size in Wuhan if the proportion of asymptomatic infections were substantial ^[8]^. In order to fill in the gap of the status of outbreak in Wuhan, we provide an estimate of the epidemic size in Wuhan based on several countries’ evacuation data.

## Methods

### Data sources

We collected information of evacuations from Wuhan using charter flights by five countries from Jan 29 to Feb 2, 2020 through their health authority website or other media. During this week, Korea has evacuated 701 people, and 2 cases were confirmed infection with an infection rate 0.29%^[9, 10]^; Japan has evacuated 565 people, and 9 cases were confirmed with an infection rate 1.59%^[11]^; Germany has evacuated 124 people, and 2 cases were confirmed with an infection rate 1.61%^[12]^; Singapore has evacuated 92 people, 4 was confirmed with an infection rate 1.08%^[13]^; France has evacuated 434 people, and none was confirmed ^[14, 15]^.

### Statistical analysis

Infection rate of 2019-nCov among people who were evacuated from Wuhan were pooled through one group meta-analysis method with random effect model. Comprehensive Meta Analysis Software V2.0 was used. Based on the number of current people stay in Wuhan, we quickly estimated the outbreak size.

## Results

Totally 1,916 people have been evacuated from Wuhan, among them 17 have been confirmed 2019-nCov infected. Based on the data above, the infection rate was calculated to be 1.1% (95% CI 0.4%-3.1%) using one group meta-analysis method with random effect model (**Figure 1**). According to the mayor of Wuhan, who introduced the progression of epidemic prevention and control of NCP, there are still more than 9 million people staying in Wuhan at that time. We estimated that there are 10 million people in Wuhan, considered the infection rate calculated above, it is estimated that almost 110,000 (95% CI: 40,000-310,000) people were infected with 2019-nCov in Wuhan around Feb 2, 2020, with an assumption that the risk of infection of evacuees is similar to Chinese in Wuhan.

**Figure 1.**
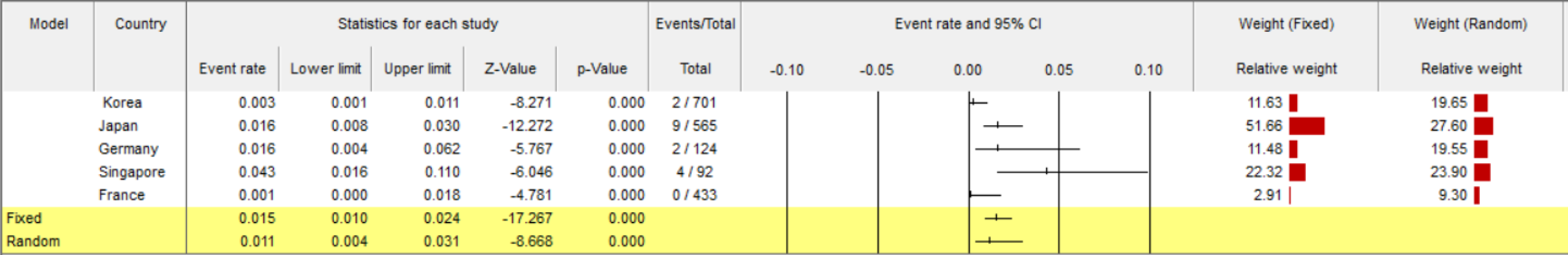
Results of one group meta-analysis for five countries’ infection rate.

## Discussion

This estimate is consistent with 21,022 (11,090-33,490) total infections from Jan 1 to 22, 2020 estimated by Read and colleagues ^[7]^ and 75,815 individuals (95% CI 37,304–130,330) as of Jan 25, 2020 by Joseph’s team if progression of the epidemic was accounted for ^[8]^, but it is much higher than the number that Chinese authority reported. Although these evacuees cannot completely represent the overall population stay in Wuhan, they tended to have certain lifestyle which may be different with local Chinese in Wuhan, and human-to-human transmission through air travel is likely during evacuation, we recommend that the estimation results should be treated with caution. Nonetheless, we believe that our results of estimated number of 2019-nCov infections in Wuhan is close to the actual situation. At the beginning of the outbreak, incidence of NCP may be vastly underestimated because of asymptomatic infection, no timely diagnosis, inadequate supplies and low sensitiviy of nCov Kit. Our result emphasizes that 2019-nCov has proposed a huge public health threats in Wuhan. China need to respond based on current actual infections and predictions based on the estimates. Joint efforts need to be done by whole society in order to control the epidemic.

## Data Availability

All data, models, and code generated or used during the study appear in the submitted article.

